# Intensive care nurses‘ experiences with patient deaths in Germany – A qualitative interview study

**DOI:** 10.64898/2025.12.05.25341281

**Authors:** Maike Michalski, Minou Gandras, Svenja Wandke, Mareike Rutenkröger, Isabelle Scholl

**Author notes:** Correspondence to: Prof. Dr. Isabelle Scholl University Medical Center Hamburg-Eppendorf, Department of Medical Psychology Martinistraße 52, Building W26 20246 Hamburg GERMANY. shared senior authorship (Mareike Rutenkröger, Isabelle Scholl).

## Abstract

**Background:** Intensive care nurses frequently encounter patient deaths, which may manifest in emotional responses, as well as cognitive and physical reactions. Evidence suggests that this can negatively impact psychological well-being, job performance and satisfaction, and may contribute to compassion fatigue or burnout. The aim of this study was to explore intensive care nurses’ experiences with patient deaths, their coping mechanisms and perceived unmet needs in Germany.

**Methods:** A qualitative study was conducted using semi-structured phone interviews from May 2024 to July 2024. Participants were 25 intensive care nurses from Germany, who were recruited by contacting nursing management staff, via social media and through direct-in-person contact. We applied purposive sampling with a maximum variation approach. The interviews were audio recorded, transcribed verbatim and analyzed using content analysis.

**Results:** The sample consisted predominantly of women (72%) and the participants experienced an average of two patient deaths per month. The participants described both emotional and cognitive reactions regarding patient deaths (e.g. grief, relief, long-term memory of the deceased). Patient deaths that occurred suddenly, under tragic circumstances or despite intensive medical efforts were experienced as particularly stressful. Frequent confrontation with death and acting against one’s own moral beliefs were described as factors that can influence emotional distress. The effects of patient deaths on ICU nurses’ personal life included a greater appreciation of their own life and health, as well as acceptance of death as natural part of life. Peer support was identified as a valuable coping mechanism, whereas attitudes toward formal support services were mixed—ranging from a desire for greater support to skepticism regarding their usefulness.

**Conclusion:** This study highlights both negative and positive effects on ICU nurses in Germany related to patient deaths, with moral distress and circumstances of death emerging as key stressors. The results underscore the need for institutional support and a more open culture around death and dying in intensive care settings. Further research should clarify which support measures are perceived as effective and for whom, in order to develop tailored interventions to mitigate professional grief.

**Trial registration:** Open Science Framework: https://doi.org/10.17605/OSF.IO/R2YVF Registration date: 11/28/2024

## Background

In many countries, including Germany, hospitals remain the most common place of death, accounting for over 50% of all deaths [1, 2]. Consequently, healthcare professionals (HCPs) are frequently confronted with the death and dying of their patients [3]. While grief is typically associated with the loss of a loved one in one’s personal life [4], the occurrence of grief in professional settings is often overlooked. Studies revealed that HCPs can experience grief following the death of a patient [5–7], which may manifest in a variety of emotional responses, as well as cognitive and physical reactions [3, 8]. Moreover, evidence suggests that repeated exposure to patient deaths and the resulting grief can negatively impact psychological well-being, job performance and satisfaction, and may contribute to compassion fatigue or burnout among HCPs [3, 6, 14].

Patients with severe illnesses are treated in intensive care units (ICUs), where the primary aim is survival. Nevertheless, mortality rates in the ICU remain high, ranging from 10 to 29% [9], exposing ICU HCPs to frequent encounters with death. Research indicates that ICU HCPs experience considerable stress due to high workload demands, the provision of end-of-life care and the frequent encounter with ethically complex and potentially traumatic situations [10, 11]. Among ICU personnel, nurses appear to be particularly affected, exhibiting a high level of grief following a patient’s death [3]. As they spend more time in direct contact with patients than any other ICU professional group [12], nurses are more likely to witness emotional distress and suffering, which contributes to elevated stress levels [13]. Given the critical condition of ICU patients, ICU nurses are intensively involved in their care [14] and may sometimes develop strong emotional connections with both patients [15] and their loved ones [16]. This closeness may contribute to the heightened levels of grief reported by ICU nurses after a patient’s death [3, 13]. A review from 2023 exploring ICU nurses’ experiences with patient deaths highlighted their feelings of responsibility in providing care for dying and deceased patients, while also indicating that this can be a source of emotional distress. This emotional burden is influenced by several factors, such as the age of the deceased or the strength of the relationship with the patient’s relatives. Coping strategies mentioned include accepting death as a natural part of life and seeking peer support. Nevertheless, some ICU nurses expressed a desire for more formal support mechanisms [16]. Although professional grief in intensive care nursing has been acknowledged in the literature, there is a notable lack of research within European contexts—particularly in Germany. This is especially relevant given the distinct training structures and professional responsibilities of nurses in the German healthcare system [17].

Therefore, understanding the experiences of ICU nurses in Germany with patient deaths, their coping strategies, and support needs is essential. This study aims to address this gap by examining ICU nurses’ grief responses, the mechanisms they use to cope, and their perceived need for intervention.

## Methods

### Study design

We conducted a qualitative interview study guided by the Criteria for Reporting Qualitative Research (COREQ) [18].

### Recruitment and sampling

This study included ICU nurses from Germany who were working or had previously worked in adult ICUs and who have experienced the death of at least one patient. To recruit participants, the study invitation was distributed via email to nursing management staff of all university hospitals in Germany, as well as to a random sample of hospitals, with a request to forward the invitation to their respective ICU nursing teams. The first author (MM) randomly selected 120 hospitals using the DIVI Intensive Care Register [19] as a basis, ensuring representation across all 16 German federal states.

The selection process considered geographic diversity (urban and rural areas), varying hospital funding (e.g., public, private, faith-based), and different levels of care provision. Recruitment efforts also included social media posts on LinkedIn and X, as well as, direct in-person recruitment by MM in her professional role as an ICU nurse. The study invitation was also shared with a professor of nursing science at HAW Hamburg for further dissemination. Interested study participants contacted the first author via email to receive further study information and schedule a telephone interview.

In order to represent a wide range of experiences we adopted a purposive sampling with a maximum variation approach regarding gender, work experience, work setting and medical specialization. The estimated sample size was determined by pragmatic saturation based on the research team’s experience with qualitative methodologies and the sample sizes used in similar studies [20]. Saturation was anticipated after approximately 25 interviews. One individual withdrew after initial email contact and did not respond to further communication; the reason for withdrawal remains unknown. Ultimately, there were 62 interested participants, which exceeded the number required to achieve the targeted sample size.

### Interviews

The first author, a female ICU nurse and medical student with no prior experience in qualitative research, conducted all one-time telephone interviews in German between May 2024 and July 2024. Interviews were audio-recorded without field notes. A pilot interview with a nurse was conducted to test the comprehensibility of the interview guide and the technical setup. This pilot interview was excluded from analysis. A research office at the University Medical Center Hamburg-Eppendorf served as private location for the interviews. Participants were instructed to choose a quiet and private setting for the interview. All semi-structured interviews were conducted using an interview guideline adapted from a previous study by the same research team [21]. The complete interview guide is available in supplementary file 1. At the beginning of each interview, participants were asked to recall and, if willing, briefly describe the death of a patient they had cared for and that had an impact on them. Subsequently, questions were asked about the experiences and impact of patient deaths, as well as, the coping mechanisms and needs of ICU nurses. Informed consent was obtained through signed forms, and participants completed a demographic and workplace questionnaire, which was submitted by post before the interview. MM had no prior relationship with any participant and clarified that the study was part of her doctoral thesis.

### Data analysis

Interviews were transcribed verbatim and imported into MAXQDA [22]. Initial transcripts were generated using the WhisperSpeaker [23] (a program integrating several open source libraries, including Whisper from OpenAI for speech recognition and pyannote.audio for speaker diarization) and then manually verified by MM and MG (co-author, medical student and research assistant). All transcripts were pseudonymized by removing all names and identifying features. Data analysis was conducted in German, with selected quotations translated into English for publication.

A combined deductive-inductive approach for content analysis was conducted following a predefined and pre-agreed analysis plan. A code system, previously developed by the same research team while conducting a similar study on professional grief among psycho-oncologists [21], served as the basis for coding this study’s material and was adjusted during the process. The first transcript was independently coded by MM and SW (PhD candidate with experience in the field of research), followed by a review meeting. The remaining transcripts were coded by MM, with a second coder reviewing and revising the codes. The second coder for the first 8 interviews was SW, and for the remaining interviews, MG. Any discrepancies were resolved via discussion, and if appropriate, the existing coding system was revised accordingly. Additionally, four meetings were held to discuss the code system in the broader research team (IS, MR, SW, MG). No participant feedback on transcripts or results was obtained.

## Results

### Sample characteristics

All N=25 participants completed the demographic questionnaire. The sample consisted of 72% women and 28% men, with the majority under 40 years of age 72% and not in leadership positions 72%. Participants worked across various ICU settings and covered different medical specialties. Detailed sample characteristics are presented in Table 1. On average, participants cared for 40 patients per month (range: 7–180) and experienced 2.3 patient deaths per month (range: 0.5–6). An average of 30% of these deaths were reported as distressing (range: 0–80%). Interviews lasted an average of 31.5 minutes (range: 22–46 minutes).

**Table 1.** Sample characteristics.

| Characteristics | Frequency (Percentage) |
| --- | --- |
| Age group |  |
| < 30 | 11 (44%) |
| 31-40 | 7 (28%) |
| 41-50 | 5 (20%) |
| 51-60 | 2 (8%) |
| > 60 | 0 (0%) |
| Gender |  |
| Female | 18 (72%) |
| Male | 7 (28%) |
| Work setting<sup>a</sup> |  |
| University hospital | 10 (31%) |
| Tertiary care hospital | 8 (25%) |
| Primary or secondary care hospital | 7 (22%) |
| Privately funded hospital | 3 (9%) |
| Faith-based hospital | 4 (13%) |
| Medical discipline<sup>a</sup> |  |
| Interdisciplinary ICU | 14 (47%) |
| Surgical ICU | 8 (27%) |
| Medical ICU | 4 (13%) |
| Early Intensive Care Rehabilitation | 1 (3%) |
| Other | 3 (10%) |
| Working experience (in years) |  |
| < 5 | 7 (28%) |
| 5-10 | 9 (36%) |
| 11-20 | 4 (16%) |
| > 20 | 4 (16%) |
| Leadership position |  |
| Yes | 7 (28%) |
| No | 18 (72%) |

<sup>a</sup>Multiple selections were possible.

### Themes and subthemes

The results of the study were categorized according to the research questions of the paper: (1) exploring the impact of patient deaths on personal and professional life of ICU nurses, (2) identifying their coping strategies and (3) their possibly unmet needs. The complete coding tree is available in supplementary file 2.

### Impact of patient deaths on ICU nurses

The impact of patient deaths varied among participants. While most reported emotional or professional effects, there were also a few who felt no or little effects. For some, patient deaths were perceived as part of professional life, with no influence on their personal life. In some interviews, participants initially did not spontaneously recognize or articulate their own emotional reactions to patient deaths, instead shifting the focus to patients’ suffering or the grief of relatives; their personal distress was described only after additional prompting. Five subthemes were identified: (1) emotional und cognitive effects, (2) influencing factors, (3) impact of the Covid-19-Pandemic, (4) impact on personal life and (5) impact on professional life.

### Emotional and cognitive effects

Common reported feelings included sadness, helplessness, and anxiety about one’s own mortality. Some reported grief, which was experienced less intensely compared to personal bereavement, due to the more distant relationship with patients. Feelings of anger were expressed in response to perceived systemic failures within the healthcare system. These emotions also arose when life-prolonging treatments were continued despite a foreseeable death, resulting in what was perceived as avoidable patient suffering. As one participant stated: “The [patient] was then ventilated for another ten hours and ultimately died. And that kind of thing just makes me angry […] and makes me sad, because people have to suffer unnecessarily.”(P04, <30). A few experienced shock, guilt, or shame, especially when a patient suddenly died or under tragic circumstances or when their absence (e.g., due to sick leave) made them feel they had let colleagues down.

Despite these unpleasant emotions, many also described feelings of contentment or relief—often linked to the perception of having provided high quality end-of-life care or the belief that the patient or relatives were no longer suffering.

At the beginning of the interviews, participants were asked to recall a personally affecting case. Nearly all were able to recall such cases, often in vivid detail, even if they occurred years or decades ago. Another cognitive phenomenon described by the participants was recurrent thoughts about deceased patients, either in their private lives or when confronted with similar patient cases.

### Influencing factors

Patient deaths are not uniformly distressing for ICU nurses, as one participant stated: “There are cases that are truly explosive, that stick with you and stay with you. And then there are others where you think, okay, it was okay that he died.” (P07, 51-60). Participants identified several factors that influence their experienced emotional stress. Most commonly, participants reported that the nature of the patient’s death significantly affected their stress levels. One participant noted: “There are two ways of experiencing death. One is when it is foreseeable […]. The other is when it happens unexpectedly in an emergency, when resuscitation or other life-saving measures fail. For me, the latter is worse, because the former is easier to come to terms with.” (P10, <30). Deaths that were sudden, unexpected, or occurred despite intensive medical efforts (e.g. after long resuscitation) or under tragic circumstances—such as after prolonged suffering or poor symptom control—were described as particularly distressing. Frequent exposure to patient deaths also contributed to cumulative emotional burden. Nurses experienced increased distress when they felt unable to provide care aligned with their professional or ethical standards. This was often attributed to systemic constraints such as time pressure and high workload, as well as situations where they had to support medical or family decisions they did not personally agree with. As one participant put it: “[…] we had to escalate the therapy at the end of the night. […] And that was difficult for me, simply because I actually wanted to minimize therapy, but the doctor didn’t really want to go along with that.” (P17, <30). Interactions with grieving relatives further intensified stress, especially when a close bond had been developed. Some nurses described offering emotional support or even crying with family members, while others felt unprepared and overwhelmed in such encounters. Additional stressors included the death of younger patients—especially those younger than the nurse—or when personal identification with the patient was high, for example, due to shared background or profession.

### Impact of the Covid-19-Pandemic

In our interview, we asked about changes in experiences with patient deaths during the COVID-19 pandemic. Some participants even reported their experiences before we asked the specific question. While some nurses saw no major change, others reported that pre-existing stressors were exacerbated. Increased frequency of deaths, more frequent resuscitations, poor working conditions, and burden caused by acting against one’s own moral standards were described as intensified during the pandemic. Restrictions on visitors were perceived as particularly difficult, as one participant stated: “Visiting opportunities were very limited, which was terrible for the patients and their relatives, but of course it also meant extra work for us, because we wanted to give the patients a certain amount of care and closeness in their final moments. It was difficult to organize our time management […].” (P15, 31-40). Deaths from COVID-19 were often perceived as prolonged and distressing. One nurse described it as “a death in instalments” (P03, 41-50). The management of the deceased—such as placing bodies in sealed bags—was also emotionally challenging. As one participant noted: “I think if you’re put in a bag and the bag is never opened again and you can’t say goodbye […], that’s terrible” (P06, 41-50). Feelings of helplessness and frustration were frequently mentioned, particularly in the early phase of the pandemic, when treatment protocols were still evolving and outcomes remained uncertain despite aggressive interventions.

### Impact on professional life

Most participants reported that they developed coping mechanisms over time due to repeated exposure to patient deaths. As one ICU nurse stated: “[…] You mature with age and experience […] Of course, this is also a learning process” (P03, 41-50). Some described a shift in perspective, recognizing death as a natural part of life and rejecting the notion that life must always be preserved at any cost. This shift often led to a greater emphasis on providing dignified, compassionate care for patients and their families. A few nurses responded to this evolving perspective by pursuing further education or advanced training. However, others experienced self-doubt regarding their professional competence. For a few, the emotional toll led to reducing work hours, transferring to different wards, or leaving the profession entirely. Some participants also noted a decline in empathy—either in themselves or observed in colleagues. While some viewed this emotional distancing as self-protective, others expressed concern about growing indifference in ICU settings.

### Impact on personal life

In their private lives, many participants reported positive effects from working closely with death. Common responses included a heightened appreciation for life and health, greater humility, and a conscious effort to prioritize meaningful life choices. Some reported lifestyle changes and initiated conversations about end-of-life wishes with loved ones, including drafting advance directives. By accepting death as part of life, few ICU nurses also reported heaving more composure in relation to their own death and the death of loved ones: “[…] because you witnessed so many deaths […], it has become a bit more normal, […]. It is still sad, it is still dramatic […], but it has somehow become more acceptable that death is part of life, […].”(P08, <30). Some participants emphasized the importance of a peaceful, pain-free, and self-determined death—not only for patients but also for themselves and their families.

Despite these positive effects, negative consequences were also reported. Many participants had difficulties separating work from private life: “There are so many things that I take home with me. The suffering of the patients […] and now I’m working hard on myself to just leave certain things in my locker [at work].” (P07, 51-60). Some experienced sleep disturbances, and a few reported psychological distress severe enough to require psychotherapeutic support.

### Coping strategies

The participants described a variety of coping mechanisms. For many, the act of caring for dying patients was itself a central form of coping —particularly when they felt adequately prepared and able to provide dignified end-of-life care. As one nurse stated: “Caring for the dying patient, particularly in their final moments, always means a great deal to me. As a result, I am better able to cope with death itself.” (P01, 31-40). Additional helpful practices included saying farewell to the deceased, preparing the body and environment for relatives and engaging in personal rituals such as prayers, opening a window or mentally honoring the deceased.

The additional coping mechanisms could be categorized into intrapersonal and interpersonal processes, as outlined in Table 2. Intrapersonal mechanisms include activities, adjustment of personal attitudes towards death, and efforts to find the right balance following the death of patients. Interpersonal coping focused on communication. In particular, the nursing team represented a central resource for both informal and formal support, such as structured debriefing of deaths during supervision sessions or case discussions.

**Table 2.**
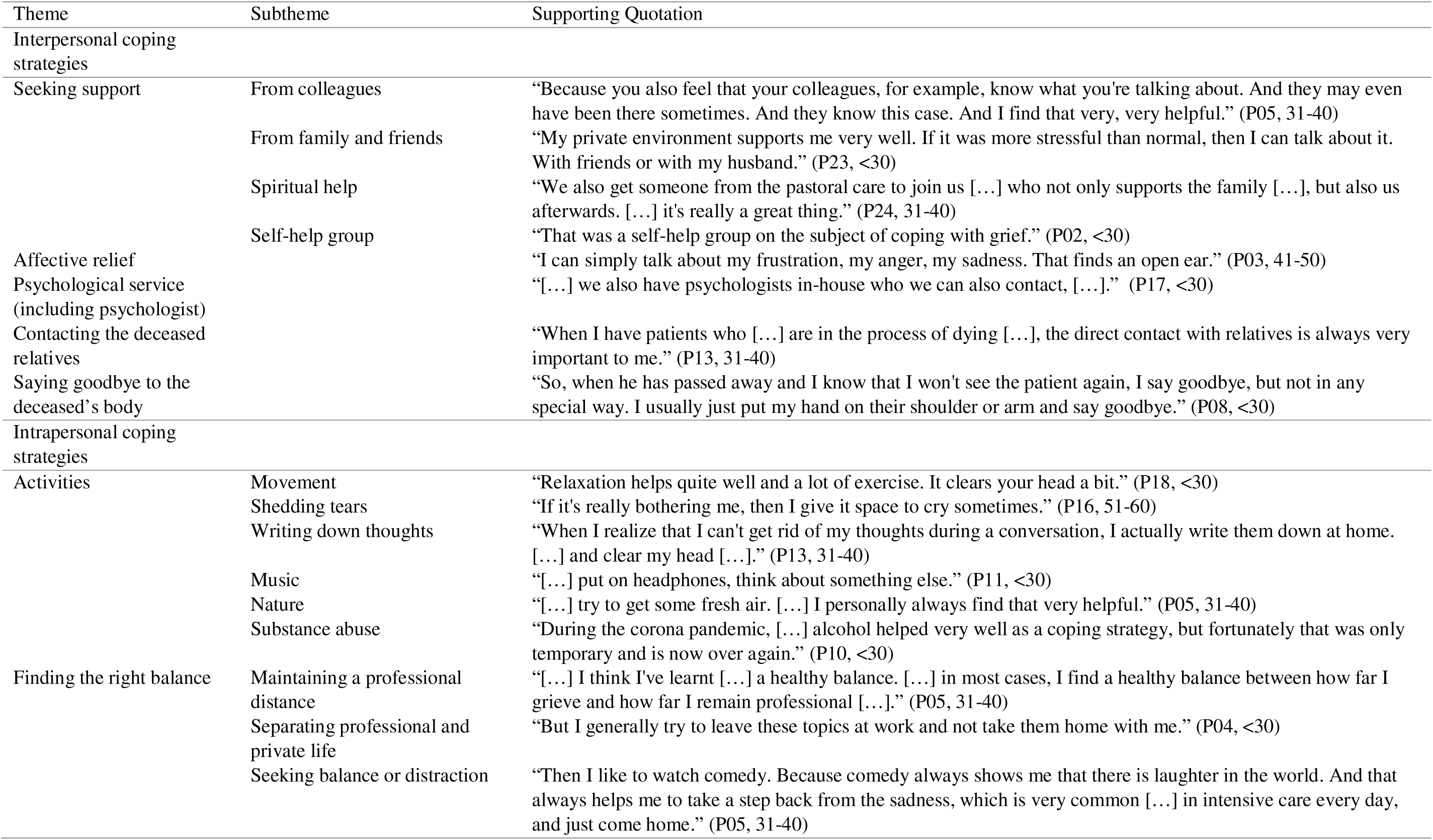
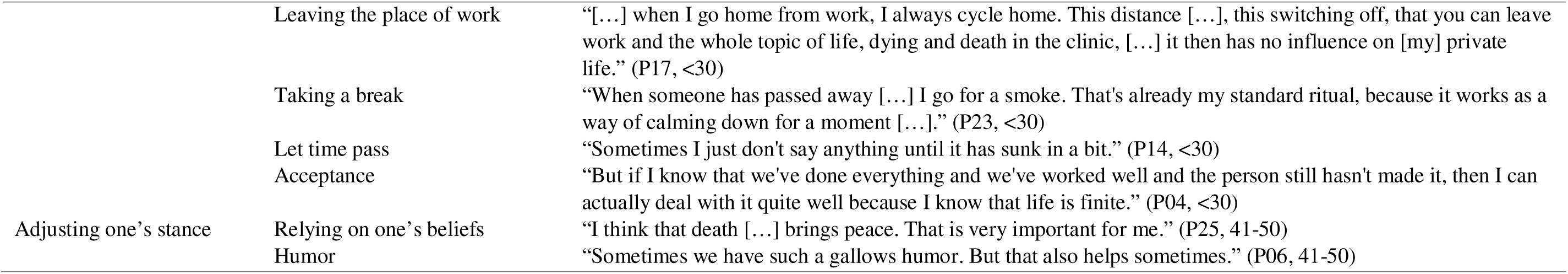
ICU Nurses strategies in coping with patient deaths.

### Barriers to successful coping

Despite the availability of coping strategies, participants identified several barriers to successfully coping with patient deaths. A recurring theme was a perceived taboo surrounding death and grief— both socially and within ICU settings. Several participants noted that death is rarely discussed openly with patients or relatives, and expressions of grief by staff may be perceived as inappropriate or unprofessional.

Communication, although perceived as supportive by some participants, was described as limited by others. Some participants reported that discussing patient deaths with non-medical friends or family offered little relief due to a lack of understanding. Others described communication with colleagues as pointless because they couldn’t express their own feelings due to prevailing taboos, or because colleagues’ reactions were inappropriate or judgmental. For some participants, formal support services such as peer support or supervision were not perceived as helpful: “It was really forced: now be emotional, now discuss what’s going on inside you. That didn’t work well.” (P08, <30).

Structural barriers further complicated coping for some participants. These included a lack of opportunities for meaningful conversation, insufficient institutional support services, and time constraints that limited opportunities for reflection. As one participant expressed: “It’s just not helpful when you […] can’t even pause for a minute because you have to run straight to the next emergency […]. I find that difficult. Then it sticks with me for longer, I think.” (P23, <30). For some participants, this situation resulted in the repression of their experience and the associated emotions.

### Unmet needs

About half of the participants reported no specific unmet needs, citing either adequate existing support structures or a lack of emotional distress related to patient deaths. In contrast, the remaining participants expressed a desire for more open dialogue about death and personal experiences with dying patients. As one nurse stated: “I sometimes wish that on our ward you could simply say […], it is and remains terrible that patients routinely lose their lives.” (P22, <30).

Some participants wished for access to specific support services, including spiritual care and individual psychological counselling. Team-based mechanisms such as regular supervision, case discussions, or farewell rituals were also seen as valuable for collective coping. Importantly, these services were expected to be offered promptly and made easily accessible.

A further need identified was education and training related to death and dying. Several participants expressed interest in workshops on grief, palliative concepts, communication with bereaved relatives, and cross-cultural or religious practices at the end of life. Although most nurses felt somewhat prepared for the topics of death and dying through prior training, many emphasized the need for improved education—particularly during onboarding in the ICU setting.

## Discussion

This study provides detailed insights into the experiences of ICU nurses in Germany confronted with patient deaths, contributing to the understanding of professional grief in this context. The findings indicate that patient deaths are not uniformly distressing for ICU nurses. Factors, such as the circumstances of death and the ICU nurse’s ability to provide end-of-life care in line with own standards, seem to influence the level of distress experienced by ICU nurses. Reported feelings included sadness and grief, emotions present in both professional and personal domains, though typically less intense and of shorter duration when related to patients. Previous studies have reported similar emotional responses among ICU nurses following patient deaths [15, 24, 25]. A novel finding identified in this study was the expression of fear regarding one’s own mortality reported by some participants. Given the high mortality rates in the ICU and the frequent exposure to patient deaths, this observation seems unsurprising; however, it contrasts with previous research that has generally reported low levels of death anxiety among nurses caring for patients at the end of life [26]. The COVID-19 pandemic may have contributed to this, as the constant threat of becoming ill oneself or having relatives fall ill was ever-present. Previous studies indicate that ICU nurses experienced death anxiety due to the COVID-19 pandemic, encompassing fears regarding both their own mortality and the loss of loved ones [27, 28]. These fears were attributed to concerns about self-infection and transmission to family members, in addition to the increased frequency of patient deaths and the nurses’ direct involvement in resuscitation efforts [27].

Professional grief emerged as a highly individual experience, varying in intensity and psychological impact—from minimal distress to severe distress. The ICU represents an acute care environment characterized by high workload, time pressure and emotionally touching patient cases. Recognition of professional grief within this context appears essential, as previous studies have demonstrated particularly high burnout rates in ICUs [11], with end-of-life care identified as a contributing factor [29]. Furthermore, situations involving patient deaths may not only evoke professional grief but also trigger moral distress. Moral distress—arising when nurses act against their moral beliefs—has been described in the field of nursing for years [30] and is reported at high levels in acute care settings such as ICUs [31]. Our study revealed that the primary burden for nursing staff was often not the patient’s death itself, but the conditions surrounding it—especially when care conflicted with individual moral values, such as in perceived overtreatment and prolonged suffering of patients. Involvement in non-beneficial interventions is a recognized contributor to moral distress, potentially leading to diminished care quality and emotional exhaustion [32].

### Implications

Our study contributes to the growing body of research on professional grief. However, further investigations are required to determine its prevalence, associated symptoms, and potential negative consequences, which may impact nurses’ well-being and job satisfaction as well as the quality of care they provide.

Many participants reported a persistent social and professional taboo surrounding death and dying in intensive care settings. This aligns with previous findings suggesting that death is often viewed merely as a routine part of the job, limiting open acknowledgement of grief [7]. To counter this, a cultural shift in ICUs towards greater openness in dealing with end-of-life issues appears necessary, allowing nurses to express emotional responses more freely.

Informal coping strategies, particularly peer support, were rated as highly effective, while attitudes toward formal support services, such as supervision, varied. Some participants desired more structured support, while others viewed it as less beneficial. Previous literature has observed a discrepancy between the proven effectiveness of such support services and their actual use among ICU nurses [33]. Further research in the German healthcare context is needed to identify which support measures are perceived as beneficial, for whom they are most effective, and to examine the factors that distract nurses from utilizing them. This would enable the development of tailored interventions to mitigate the impact of professional grief. Additionally, institutions should facilitate and promote greater opportunities for peer support among caregivers.

### Strengths and limitations

To the best of our knowledge, this is the first study conducted in Germany that examines the experiences of ICU nurses regarding patient deaths. The sample consisted of ICU nurses from various specialties and hospitals across different regions of Germany. Data analysis was performed by three coders, two of them not involved in data collection. The analysis followed a predefined analysis plan and was regularly reviewed by a team with expertise in qualitative methods. The study adhered to the COREQ guidelines.

However, several limitations must be acknowledged. Due to the absence of centralized registry for nursing professionals or a unified professional body in Germany, there is no mechanism for reaching all ICU nurses nationwide. Additionally, the sample included a higher proportion of female participants, reflecting the gender distribution within the nursing profession; however, this must be considered when interpreting the findings. The use of telephone interviews represents a further limitation, as non-verbal behavior and reactions of the interviewees could not be captured. Nevertheless, this mode of data collection was deliberately chosen to reduce logistical demands on participants and to facilitate inclusion of nurses from diverse regions across Germany. Transcripts and results were not returned to participants due to the anticipated burden. Despite careful transcription by two individuals, minor errors cannot be entirely ruled out.

## Conclusion

By examining the experiences of ICU nurses in Germany with patient deaths, this study identified both professional and personal impacts, along with factors contributing to increased distress. The effects were not always negative: ICU nurses also perceived positive impacts from their experiences with patient deaths, such as a heightened appreciation for life. Moral distress and circumstances of death were identified as additional stressors. The findings provide a foundation for further research on professional grief in intensive care settings in Germany and for the development of appropriate support services for ICU staff. Future investigations should address ICU HCPs attitudes towards support services to clarify which measures are perceived as suitable and effective. Team communication emerged as a central coping strategy, highlighting the importance of institutional efforts to support and foster such exchanges actively. Nurses expressed diverse needs, ranging from enhanced institutional support to the establishment of a more open culture surrounding death and dying in intensive care. Accordingly, improving available support services and reducing the existing taboos around death and grief in intensive care appear to be necessary steps.

## Data availability

The qualitative data collected and analyzed (in German) are available from the corresponding author on reasonable request. Signing a data use/sharing agreement will be necessary, and data security regulations both in Germany and in the country of the investigator, who proposes to use the data, must be complied with. Preparing data sets for use by other investigators requires substantial work and is thus linked to available or provided resources.

## Abbreviations

ICU: Intensive care unit
HCP: Healthcare professional

## Acknowledgements

Not applicable.

## Author information

### Contributions

IS and MR are the responsible principal investigators of the study and share senior authorship. IS, MM, MG, SW and MR were involved in planning and preparation of the study. MM recruited participants and collected data, and analyzed the data with help of MG and SW. All authors interpreted the results. MM wrote the first draft of the manuscript. MG, SW, MR and IS critically revised the manuscript for important intellectual content. All authors gave final approval of the version to be published and agreed to be accountable for the work.

### Corresponding Author

Correspondence to Isabelle Scholl.

## Ethics declarations

### Ethics approval and consent to participate

This study adhered to the Helsinki Declaration of the World Medical Association and respected principles of good scientific practice. Ethical approval was obtained prior to investigation by the local Ethics Committee of the University Medical Center Hamburg-Eppendorf (approval number: LPEK- 0706). Participation was voluntary and the participants were fully informed about the purpose of the study, data collection and the use of collected data. Written informed consent was obtained from all included participants.

### Consent for publication

Not applicable.

### Supplementary information

Supplementary file 1_interview guide.docx

Supplementary file 2_coding tree.docx

