## Supplementary file 1 for "Intensive care nurses‘ experiences with patient deaths in Germany – A qualitative interview study"

**Supplementary file 1: Interview Guide**

| **Verbal consent** | We will now proceed and start our interview. I will switch on the recording devices.  (MM switches on devices)  I have now started the audio recording, is that okay with you?  (Verbal consent?)  Then let's start now. |
| --- | --- |
| **Subject** | **Probing questions** |
| Introduction  Sharing own experiences (leading over to the experience of professional grief) | In our interview today, I would like to ask you about your personal experiences with the death of patients in the intensive care unit. It will also be about your coping mechanisms. There is no right or wrong here; I am interested in your individual perspectives.  I would like to ask you to remember a case in which a patient you cared for died in the intensive care unit? If you like, you can tell me briefly about this case. |
| Experiences with patient death | 1. How did you experience the death you just described to me, or other patient deaths?    a. How would you describe the feelings and thoughts after your patient has passed away?    b. How have these feelings developed/changed over time?    c. If you had to give a name to your reactions to the death of a patient, how would you call them?    d. How have your experiences with the death of patients changed during the Covid-19 pandemic? 2. Now let’s talk about the topic of death in general: What are your personal beliefs or convictions regarding death?    a. Do you have a specific idea of a "good" death?    b. Have these changed over the course of your experiences with patient deaths? 3. To what extent does your experience with the death of this patient or of patients in general differ from your experience with private deaths?    a. Are there any similarities between the two experiences?    b. Are there any differences? |
| Impact of patient deaths | 4. How did your experiences with patient deaths affect your own life?    a. Were there any professional or private effects?    b. Can you describe these effects to me in more detail? 5. In past cases of deceased patients, how did your surroundings react when you talked about the topic?    a. In a professional context?    b. In a private context?    c. Did you wish for or hope for different reactions? |
| Coping mechanisms for professional grief | 6. How did you cope with the death of the patient you just told me about, or with the death of other patients?    a. What strategies do you use to process these experiences?    b. Are there activities you find comforting/helpful after a patient has died?    c. Which of these strategies do you find helpful in coping with the death of patients? Are there any that are not helpful for you? 7. Do you have a (farewell) ritual (personally/in your team) when a patient has passed away? 8. What advice would you give to new/future intensive care nurses who are confronted with the death of a patient? |
| Support needs in coping with patient deaths | 9. Where do you find support for coping with the death of patients when you need it?    a. Are there offers for support in your professional environment? 10. Would you wish for (additional) support services? If so, which ones? |
| (Educational) Preparation for coping with patient deaths | 11. Were you prepared for coping with the death of patients during the course of your professional training?     a. If yes, could you describe what that preparation looked like?     b. If no: how would you have liked that preparation to be?     c. Looking back, would you have wished for different preparation? Was something specific missing? 12. Would you like ongoing or further training opportunities in dealing with the death of patients?     a. In what context would you prefer such training? |
| **Ending of the interview**  Open question | 13. Finally, I would like to know if there is anything else you would like to share with me regarding your experiences with the death of patients that I may not have asked you before? |
