## Supplementary file 2 for "Intensive care nurses‘ experiences with patient deaths in Germany – A qualitative interview study"

**Supplementary file 2: Coding tree**

| Theme | Subtheme (Level 1) | Subtheme (Level 2) | Subtheme (Level 3) | Description |
| --- | --- | --- | --- | --- |
| Research question 1: How do patient deaths affect intensive care nurses? | | | | |
| Few/ no effects |  |  |  | Participants report no or few noticeable effects from one or more patient deaths; can be coded alongside emotions and/or other effects. |
| Being touched |  |  |  | Participant reports that a patient’s death/patient deaths evoke emotions; affects them emotionally, without a negative connotation. |
| Being distressed |  |  |  | Participant reports emotional distress caused by a patient’s death/patient deaths; corresponds to a negative connoted version of ‘being touched’. |
| Doubts about the legitimacy of own emotions/  distress |  |  |  | Participants question the extent to which their own (emotional) concern/reactions are appropriate; indicates disenfranchised grief. |
|  | Limited space for own emotions |  |  | Participant reports feeling as if his/her own (emotional) reaction (grief) has more space or is (societally perceived) as more appropriate in regard to deaths occurring in the private sphere. |
|  | Grief is perceived as a taboo subject |  |  | Participant reports perceiving grief/death as a taboo subject, socially or in the intensive care unit setting. Also: grief is perceived as disenfranchised in a professional context. |
| Factors contributing to higher/lower distress following patient deaths |  |  |  | Why do clinicians find some deaths more distressing than others? |
|  | Work under Covid-19 conditions (additional code) |  |  | Participant reports that the Covid-19 pandemic was an additional stress factor in dealing with patient death. This code can only be coded in connection with other stress factors. |
|  | Related to the nature of the death/manner of dying |  |  | Participant reports increased distress due to direct presence at the moment of death, especially in patients who are in pain during death or suffer from a lack of symptom control, unexpected deaths or a high frequency/density of patient deaths or the circumstances that led to death. |
|  | Related to the nurse-patient-relationship |  |  | Participants report increased distress when they are dissatisfied with their care (also due to external factors like inability to control symptoms), strong sympathy between nurse and patient, or long-term and deep relationships. |
|  | Related to the relatives of the patient |  |  | Participant reports that it is particularly stressful for him/her when he/she is confronted with the grief of relatives. |
|  | Related to patient characteristics |  |  | Participant reports increased distress when patients do not accept/deny their impending death or when patients are young (especially children). |
|  | Related to the nurses’ personal life or characteristics |  |  | Participant reports a reduced capacity for resilience or reduced emotional stability when there is a high identification with patients (e.g., similar age, professional proximity, shared migration background), during private deaths or other crises or an overall reduced capacity for resilience or emotional stability. |
|  | Care practices conflicting with moral values |  |  | Participant reports an additional burden if care of the dying was not possible in accordance with the patients’ wishes and own (moral) values (e.g. moral distress). Example: Overtreatment of very old people. |
|  | Care practices conflicting with systemic conditions |  |  | Participant reports stress due to workplace conditions, e.g. insufficient personnel resources, rules under Covid-19 pandemic. |
| Emotional impact |  |  |  |  |
|  | Content |  |  | Participant reports feelings of content, e.g. with own work, quality of provided care. |
|  | Fear of own mortality |  |  | Participant reports fear of his/her own finiteness due to the (frequent) confrontation with the death of patients or with the way in which patients die. |
|  | Relief |  |  | Participant reports feeling relieved, e.g. because the deceased patient’s suffering came to an end due to death. |
|  | Gratefulness |  |  | Participant reports feeling grateful, e.g. for shared relationship, trust and intimacy with the deceased patient. |
|  | World-weariness (“Weltschmerz”) |  |  | Participant reports feeling somewhat weary about e.g. life being unfair. |
|  | Helplessness |  |  | Participant reports feeling helpless, overwhelmed, left alone in some sort; ‘resources do not match tasks’. |
|  | Sadness |  |  | Participant reports feeling sad. |
|  | Shock |  |  | Participant reports feeling surprised, taken off-guard, shocked by patient death. |
|  | Anger |  |  | Participant reports feelings of anger, e.g. towards the deceased patient, the circumstances under which care had to be provided (component of moral distress), etc. |
|  | Guilt |  |  | Participant reports feeling guilty. |
|  | Shame |  |  | Participant reports feelings of shame surrounding patient death; e.g. due to own distress. |
|  | Grief |  |  | Participant reports feeling grief about a patient’s death. |
| Cognitive impact |  |  |  |  |
|  | Reoccurring thoughts about patient(s) |  |  | Participant reports lasting or reoccurring thoughts about deceased patient(s). |
|  | Long-term intense memories of deceased patients |  |  | Participant reports memories that can be recalled in detail in the long term, e.g. ‘as if it had happened yesterday’. |
| Impact on professional life/performance |  |  |  |  |
|  | Impaired ability to empathize with patients |  |  | Participant reports feeling less able to empathize with patients due to confrontation with patient deaths. |
|  | Doubts about professional competence |  |  | Participant reports doubting his/her own professionalism/ overall competence as an intensive care nurse; feelings of inadequacy. |
|  | Routine in dealing with patient deaths |  |  | Participant reports developing/having developed a routine in experiencing and coping with patient deaths; frequent confrontation leads to familiarization, which aids in keeping composure. |
|  | Reframing one’s professional self-concept |  |  | Participant reports that the frequent confrontation with the death of patients has led to a change in the own understanding of profession. The view of what good care for a (dying) patient looks like has changed. A great deal of effort is put into the care of patients and their relatives. |
|  | Quit job, change ward, reduction of working hours |  |  | Participant reported that he/she had quit jobs or moved to another ward or reduced his/her working hours. |
|  | Start Further education or studies |  |  | Participant reports to take up studies or further training with the aim of being able to provide better end-of-life-care or to accompany relatives accordingly. |
| Impact on personal life |  |  |  | Spill-over; effects that present in personal life as well (might be present in professional life too/’spilled-over’ from professional into private domain. |
|  | Sleep disturbances |  |  | Participant reports sleep disorder. |
|  | Consolidate/clarify beliefs concerning death |  |  | Participant reports on changes in beliefs concerning death. She/he was able to define for herself/himself what a good death means. |
|  | Utilization of psychotherapeutic support |  |  | Participant reports that he/she is/was undergoing psychotherapeutic treatment for work-related issues (including grief-related aspects). |
|  | Active engagement with one's own death |  |  | Participant reports that he/she has actively prepared for his/her own death/dying e.g. in the form of a living will or health care proxy. |
|  | Serenity in dealing with death and dying |  |  | Participant reports frequent confrontation with death/dying in professional life lead to increased serenity/composure in dealing with death of loved ones. |
|  | Difficulties separating private and professional life |  |  | Participant reports difficulties in keeping boundaries between professional and private life/ maintaining a work life balance; participant does not have to evaluate the “lack of boundaries” as negative. |
|  | Heighted appreciation of own life/ giving perspective |  |  | Participant reports exposure to patient deaths has shown him/her to value his/her own life and health (or that of relatives) and aided him/her in e.g. setting personal priorities, making decisions that are right for oneself, living life the way that is right for one personally (in the knowledge that life itself is fleeting). |
| Differences between professional grief and grief in general |  |  |  |  |
|  | Difference in the way of saying goodbye |  |  | Participant reports differences in the way they say goodbye to the deceased; e.g. attending funerals is appropriate within the private domain, but less appropriate or inappropriate concerning deceased patients. |
|  | Difference in the duration of grief |  |  | Participant reports professional grief differs in duration (taking less time). |
|  | Difference in felt intensity |  |  |  |
|  |  | Due to increased regulatory capacity in the event of patient death |  | Participant reports self-regulation and emotion regulation are easier regarding patient deaths; might be due to higher demands on one's own ability to regulate (professionalism). |
|  |  | Due to differences in the relationship’s depth |  | Participant reports differences in intensity due sharing a ‘less close bond’ with patients, private relationships by nature being deeper and therefore more distressing in the event of loss. |
|  |  | Due to grief being confined to professional role |  | Participant reports with personal losses the person as a whole is emotionally shaken, the loss is fundamental, whereas professional grief ‘only’ affects the professional role. |
| Research question 2: How do intensive care nurses cope with patient deaths? | | | | |
| Strategies for coping |  |  |  | Strategies that aid in ‘rounding off’ the relationship; help intensive care nurses to ‘let go of the patient’. |
|  | Anticipatory coping through end-of-life care |  |  | Participant reports it is helpful to provide good end-of-life care for dying patients. |
|  | Intrapersonal strategies |  |  | ‘no other person needed’ |
|  |  | Activities |  |  |
|  |  |  | Nature | Participant reports using nature/natural environment as a resource. |
|  |  |  | Music | Participant reports listening to music to cope with patient deaths. |
|  |  |  | Writing down thoughts | Participant reports writing down thoughts e.g. in a diary or in form of a poem helps in coping. |
|  |  |  | Shedding tears | Participant reports that he/she cries to express emotions. |
|  |  |  | Movement | Participant reports using movement to cope with patient deaths, e.g. going for a walk or doing sports etc. |
|  |  | Adjusting one’s stance |  |  |
|  |  |  | Relying on one’s beliefs | Participant reports relying on established beliefs to reframe patient deaths; e.g. explicit perceptions like ‘better for the patient or saved from worse’ or religious beliefs. |
|  |  |  | Acceptance | Participant reports using acceptance aids them in coping with patient deaths. Accepting death as a part of life. |
|  |  |  | Humor | Participant reports using humor as way of dealing with the death of patients. |
|  |  | Finding the right balance |  |  |
|  |  |  | Let time pass | Participant reports that it helps them to consciously allow time to pass when coping with patient deaths. |
|  |  |  | Taking a break | Participant reports taking either small breaks/moments to themselves to cope with patient deaths and/or using time off of work and vacations to cope. |
|  |  |  | Seeking balance or distraction | Participant reports distracting himself/herself or explicitly doing leisure ‘light’ leisure activities to balance out ‘heavy’ work-related events as coping strategy. |
|  |  |  | Leaving the place of work | Participant reports leaving the place of work, having ‘a way home’ as a coping strategy; distance between work and home life as helpful. |
|  |  |  | Separating professional and private life | Participant reports maintaining boundaries between work and private life as coping strategy. |
|  |  |  | Maintaining a professional distance | Participant reports that maintaining a professional distance helps them both before and after the death of their patients. The extent of professional distance depends on the subjective perception of the participant and can be both functional and dysfunctional when viewed objectively. |
|  | Interpersonal strategies |  |  | Other people have to be present in some capacity. |
|  |  | Social Support |  |  |
|  |  |  | Through colleagues | Participant reports peer support from medical or nursing colleagues as coping strategy; informal or formalized (through supervision or peer counselling groups). |
|  |  |  | Through friends and family | Participant reports sharing a patient’s death with partners, friends or family as a coping strategy. |
|  |  |  | Through spiritual assistance | Participant reports seeking spiritual guidance or talking to e.g. clergy as a coping strategy. |
|  |  |  | Through self-help groups | Participant reports attending self-help groups as a coping strategy. |
|  |  | Psychological service/psychologist |  | Participant reports talking to psychologists or using psychological services as coping strategies. |
|  |  | Affective relief |  | Participant reports showing and/or sharing own emotions helps him/her in coping with patient deaths. |
|  |  | Saying goodbye to the deceased’s body |  | Participant reports (re)visiting the body of the deceased aids them in coping. |
|  |  | Contacting the deceased relatives |  | Participant reports contacting the deceased’s relatives helps him/her in coping with the patient’s death. |
|  | Rituals |  |  | Any coping strategy performed in a ritualized manner, usually repetitive. |
|  |  | Prayer |  | Participant reports that practicing his/her religion in the form of prayer helps him/her cope with the death of patients. |
|  |  | Paying tribute to the deceased |  | Participant reports paying tribute in some way as important, a component of individually chosen ritual. |
|  |  | Open a window |  | Participant reports opening the window following a patient’s death, e.g. in the belief to allow the soul to escape. |
|  |  | Preparation of the deceased and/or the surroundings |  | Participant reports that it is important for him/her to provide final care for the deceased and to arrange the surroundings in a pleasant manner. |
|  | Substance abuse |  |  | Participant reports using drugs to distract themselves from the death of patients or to self-medicate negative stressors (e.g. sleep disturbances). |
|  | Continue with the work |  |  | Participant reports that in the event of a patient's death, he/she continues working in order to cope with it. |
| Barriers in coping with patient deaths |  |  |  | Everything hindering psycho-oncologists in successfully coping with patient deaths/ ‘rounding off’ the relationship. |
|  | Expression of grief is impossible |  |  | Participant reports expression of own emotions, distress or grief is or feels impossible, e.g. due to interlocutors being inexistent or unavailable. |
|  | Time constraints |  |  | Participant reports feeling like he/she does not have enough time to feel and express their own emotions or to say goodbye, e.g. due to hectic schedule, other task’s urgency etc. |
|  | Pressure/perfectionism in coping with patient deaths |  |  | Participant reports own expectation or feeling pressured by other’s expectations; that there is one right way to deal with the death of patients. |
|  | Suppression of emotions |  |  | Participant reports ‘not wanting to feel’ his/her own emotions/grief, e.g. avoiding situations that ‘trigger’ grief or cognitively suppressing one's own emotions. |
|  | No (institutional) support services available |  |  | Participant reports that there are no support services available in his professional environment in the event of a patient's death, or that he is not aware of any such services. |
|  | Sharing experiences of patient deaths is perceived unhelpful |  |  |  |
|  |  | With friends and family |  | Participant reports that sharing experiences of patient deaths with friends and family are perceived as unhelpful. Possible reasons for this include a lack of understanding of the interlocutor or misunderstandings. |
|  |  | With colleagues |  | Participant reports that sharing experiences of patient deaths with colleagues are perceived as unhelpful. Possible reasons for this include a lack of understanding of the interlocutor or the occurrence of blame within the team. |
| Research question 3: Were intensive care nurses educationally prepared to cope with patient deaths? And if so: where? | | | | |
| YES, pre-graduate education |  |  |  | Participant reports being prepared to cope with the death of patients as part of his/her nursing training. |
|  | Theoretical |  |  | Participant reports preparation as part of theoretical instructions in nursing school. |
|  | Through practical experiences |  |  | Participant reports preparation as part practical assignments at the hospital. |
| YES, special training for intensive care nursing |  |  |  | Participant reports being prepared to cope with patient deaths as part of their special training to become an intensive care specialist. |
| YES, nursing studies |  |  |  | Participant reports being prepared to cope with patient deaths as part of his/her nursing studies. |
| YES, post-graduate training |  |  |  | Participant reports being prepared to cope with patient deaths in post-graduate training. |
| NO, pre-graduate training |  |  |  | Participant reports not being prepared to cope with patient deaths or feeling like preparation was insufficient within education in nursing training. |
|  | Theoretical |  |  | Participant reports not being prepared to cope with patient deaths or feeling like preparation was insufficient within theoretical instructions in nursing school. |
|  | Through practical experiences |  |  | Participant reports not being prepared to cope with patient deaths or feeling like preparation was insufficient within practical assignments at the hospital. |
| NO, special training for intensive care nursing |  |  |  | Participant reports not being prepared to cope with patient deaths or feeling like preparation was insufficient within special training to become an intensive care specialist. |
| Research question 4: Do intensive care nurses have any unmet needs in regard to their coping with patient deaths? | | | | |
| No unmet needs |  |  |  | Participant reports personally not having any unmet needs in regard to patient deaths. |
| Prompt availability of support services |  |  |  | Participant reports that it would be helpful to receive support shortly after the death of a patient. |
| Space to discuss death and own emotional distress |  |  |  | Participant reports wishing for more space to discuss death and share own emotions in regard to patient deaths. |
| Within a team |  |  |  |  |
|  | Case discussions |  |  | Participant reports wishing for more case discussions involving deceased patients to facilitate professional reflection. |
|  | Supervision |  |  | Participant wishes for more (frequent) team supervision. |
|  | Team-based rituals |  |  | Participant shares a desire to take part in team-based rituals to say goodbye to deceased patients. |
| Individually |  |  |  |  |
|  | One-on-one conversations |  |  | Participant shares a need for more opportunities to have one-on-one conversations concerning his/her distress with patient deaths. |
|  | One-on-one supervision |  |  | Participant reports needing more opportunities for supervision, WITHOUT other team members present. |
| Educational and informational needs |  |  |  | Participant reports needing additional education and/or information surrounding patient deaths, coping with them etc. |
|  | Death/ bereavement workshops |  |  | Participant wishes for workshops surrounding death or bereavement. |
|  | Training on dealing with relatives |  |  | Participant wishes for further training on dealing with relatives of deceased patients. |
|  | Information on palliative care concepts |  |  | The participant wishes more information on dealing with dying patients and on palliative care concepts. |
